# The impact of repeated rapid test strategies on the effectiveness of at-home antiviral treatments for SARS-CoV-2

**DOI:** 10.1101/2022.01.18.22269408

**Authors:** Tigist F Menkir, Christl A Donnelly

## Abstract

As has been consistently demonstrated, rapid tests administered at regular intervals can offer significant benefits to both individuals and their communities at large by helping identify whether an individual is infected and potentially infectious. An additional advantage to the tested individuals is that positive tests may be provided sufficiently early enough during their infections that treatment with antiviral treatments can effectively inhibit development of severe disease, particularly when PCR uptake is limited and/or delays to receipt of results are substantial. Here, we provide a quantitative illustration of the extent to which rapid tests administered at various intervals can deliver benefits accrued from the novel Pfizer treatment (nirmatrelvir) among high-risk populations. We find that strategies in which tests are administered more frequently, i.e. every other day or every three days, are associated with greater reductions in the risk of hospitalization with weighted risk ratios ranging from 0.17 (95% CI: 0.11-0.28) to 0.77 (95% CI: 0.69-0.83) and correspondingly, higher proportions of the infected population benefiting from treatment, ranging from 0.26 (95% CI: 0.18-0.34) to 0.92 (95% CI: 0.80-0.98). We further observed that reduced positive-test-to-treatment delays and increased testing and treatment coverage have a critical influence on average treatment benefits, confirming the significance of access.

## Main text

Rapid tests for SARS-CoV-2 have been shown to help identify individuals who may be infectious.^1–3^ Their newfound use, particularly among those prone to severe disease, is identifying infections early enough that they can be effectively treated with antiviral treatments, including the Pfizer drug PF-07321332 (nirmatrelvir)^4,5^, which necessitates early use to lower the risk of hospitalization. Here, we demonstrate that testing rates, as well as testing and treatment coverage and positive-test-to-treatment delays, shape the impacts of such test-and-treat policies.

Many have promoted rapid testing to identify infections when antivirals are still helpful^6–8^, so there is a need to quantify the extent to which frequent rapid testing can enable high-risk patients to benefit most from the treatment. We thus build on prior studies which characterized the ability of rapid testing strategies to identify presymptomatic patients or to reduce transmission.^9–13^ Specifically, acknowledging the short window over which treatment can effectively inhibit more severe outcomes, we assess different strategies - defined by varying rates of test administration - in their relative ability to curtail the risk of hospitalization among an adult patient population facing an increased risk of severe disease, i.e. those who would be offered treatment in the event of testing positive.

To evaluate the benefits of repeated rapid testing at different rates on treatment effects, we used inferred lateral flow test (LFT)-associated positivity estimates from a Hellewell et al. analysis^9^ and estimated hospitalization risks when treated within three and five days following the onset of symptoms from the latest summary of the Phase 2/3 EPIC-HR trial findings.^4^ Specifically, for each rapid testing strategy (every other day, every three days, once a week, once every two weeks, strategies explored in Larremore et al.^10^, and once only after symptom onset) we estimated test-positivity-probability-weighted risk ratios (RRs) of hospitalization – hereafter referred to as ‘weighted RRs of hospitalization’ – as a function of time since infection, the proportion of the infected population who would be offered the treatment, and the proportion of the infected population who would take it sufficiently early to benefit from treatment. In sum, to generate weighted RRs for each testing regime, we assigned probabilities for every possible testing sequence consistent with the regime, leveraging the Hellewell et al. positivity estimates^9^ as a function of time since infection, to period-specific ratios comparing the risk of hospitalization in treatment and placebo groups, leveraging the EPIC-HR summary data.

To estimate the proportion of the infected population offered treatment under each testing regime, we again used the Hellewell et al. positivity estimates to yield the complement of the proportion of the population never testing positive over all possible testing sequences. Additionally, we estimated the proportion of the population who would be given treatment at a time when it is associated with a non-zero reduction in the risk of hospitalization in the same way we generated weighted RRs, although instead weighting indicators of whether the test is conducted during the clinically relevant window. We further evaluated the proportion of the infected population offered/benefiting from treatment under a one-time testing strategy immediately following symptom onset. Finally, we explored the sensitivity of our findings to assumed treatment efficacy trends, an incubation period distribution more consistent with Omicron infections^14^, and three measures of access: treatment uptake or coverage, the delay from testing positive to treatment, and testing coverage.

As expected, we found that when tests are administered more frequently, the benefits associated with nirmatrelvir initiation increase dramatically, such that treatment substantially reduces the risk of hospitalization (Figure 1A). While the median RR associated with the every other day strategy is 0.17 (95% CI: 0.11-0.28), the median RR associated with the once every two weeks strategy is 0.77 (95% CI: 0.69-0.83), with a dramatic increase in median RRs from the two higher-frequency testing regimes to the less-frequent testing alternatives (Figure 1A). Correspondingly, we see a pronounced increase in the proportion of the infected population benefiting from treatment as testing frequency increases, ranging from 0.26 (95% CI: 0.19-0.34) to 0.92 (95% CI: 0.80-0.98) (Figure 1B). The estimates of proportion given the treatment and proportion actually deriving some benefit from it indicate that nearly everyone who tests positive and thus takes treatment receives some benefit. This arises because, in the estimated Hellewell et al. positivity curves^9^, nearly all positive tests occur within two weeks of infection. Consequently, under our base case scenario, where drug-associated benefits extend to seven days since symptom onset (which corresponds to twelve days since infection assuming an incubation period of five days), almost all individuals who test positive are captured within this drug efficacy window.

**Figure 1.**
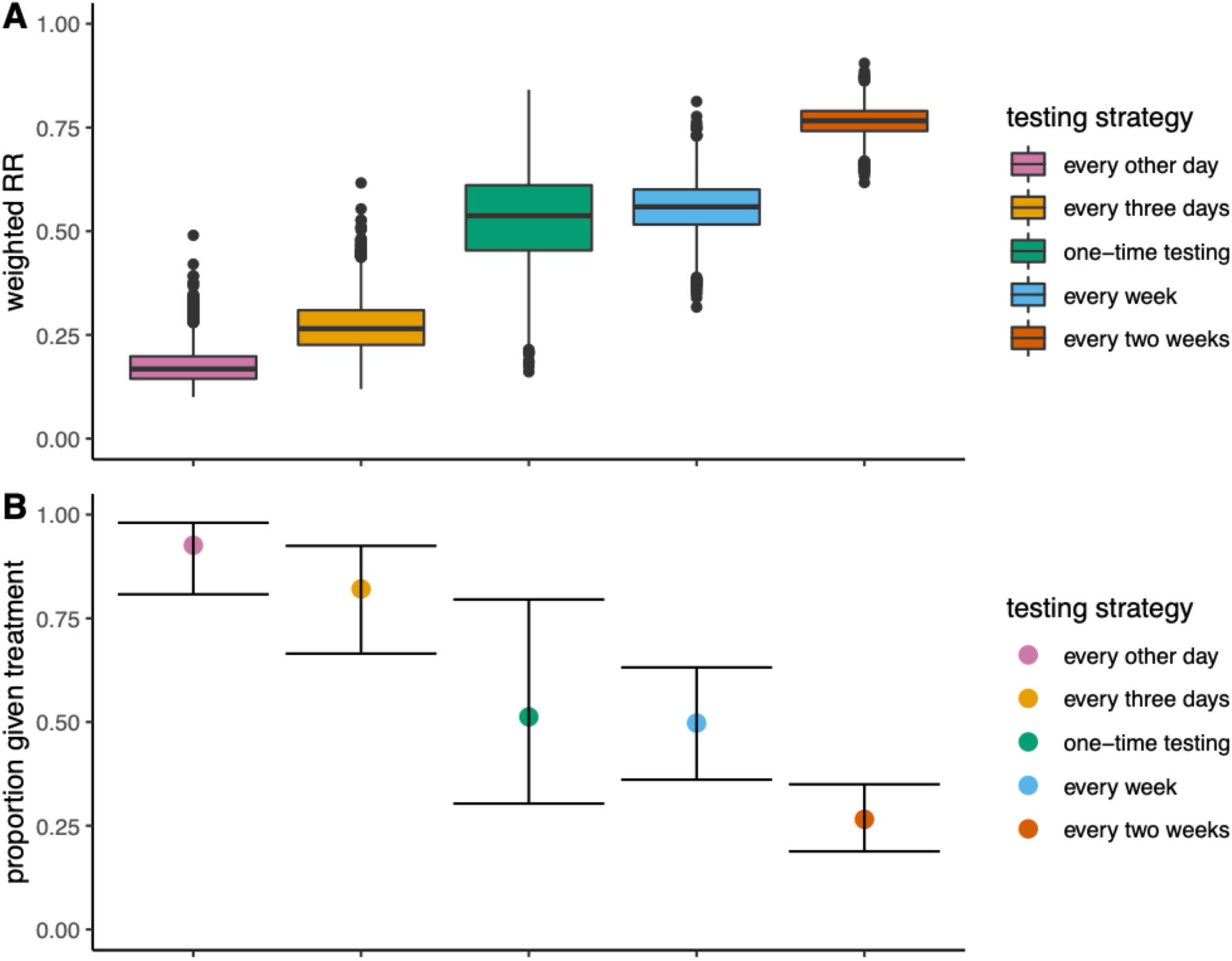
(A) Distribution of estimated weighted RRs of hospitalization by testing strategy: every other day, every three days, one-time testing, every week, and every two weeks. Medians are marked by solid horizontal lines, each box includes the full interquartile range, and plotted points are those which extend beyond the upper/lower quartile +/- 1.5*interquartile range (B) Estimated proportions given treatment by testing strategy (including the one-time post-symptom onset testing strategy) with 95% uncertainty intervals. In all cases no positive-test-to-treatment delay and full test coverage were assumed.

Under a sensitivity analysis in which we assume a distribution of shorter incubation times, to reflect time-to-symptom-onset trends for patients infected with the Omicron variant, our estimates indicate that fewer individuals are able to benefit from treatment (Supplementary Table S1), as expected when symptoms develop more quickly and there is a reduced opportunity to test when treatment is more effective. However, we note that the differences in estimated proportions benefited between the two scenarios are modest (Supplementary Table S1).

In comparison to the multi-frequency testing strategies, an approach of testing once after symptoms arise results in a notable proportion of the infected population given treatment, but with substantial variability (0.51, 95% CI: (0.30-0.80) and 0.42, 95% CI: (0.080-0.80) for our baseline scenario and shorter incubation period scenario, respectively) (Figures 1 and S2). Importantly, while this strategy - for the baseline scenario - was found to outcompete the lower-frequency strategies of testing every week and every two weeks, its expected impact is far eclipsed by the every-other-day and every-three-days strategies, with only the latter two enabling a strong majority of the population to be offered treatment. We note that under the shorter incubation scenario, however, the one-time testing strategy instead reports a lower proportion offered treatment than the once-every-week strategy. Our baseline results highlight the essential trade-offs between testing costs and treatment impacts; despite the increased investment that would be required for more-frequent testing, a vastly increased proportion of the at-risk population would be afforded the opportunity to benefit from treatment. Furthermore, that the weekly and bi-weekly testing regimes are generally less effective than simply testing once symptoms emerge highlights that more-frequent testing is essential for any repeated testing policy to have any real added treatment-associated benefits. These findings replicate what have been observed in prior studies about the crucial role of “test frequency” in the transmission-limiting context^10,11,13^, suggesting that such regular testing regimes would assure two-fold benefits in infection prevention and disease control. A potential hybrid testing scheme might also be worth considering, with repeated testing as well as a test immediately following the onset of symptoms, should they occur, which would provide some intermediate benefit at some intermediate cost than its more or less frequent testing counterparts.

We found that treatment benefits depend on both treatment and test coverage and the delay from testing positive to treatment (Figure 3). To achieve RRs within the range of what we observed with full coverage, zero delays and testing every other day, treatment coverage of at least 70% would require positive-test-to-treatment delays of no more than two days. With more sparse testing, treatment coverage and positive-test-to-treatment delays are critical, with smaller RRs achieved only through nearly full coverage and delays of no more than two days. When we independently assess the impacts of testing coverage, we find that estimated proportions benefiting from treatment are particularly sensitive to the assumed proportion testing, particularly for the more-frequent-testing strategies (Supplementary Figures S3). However, we find that when we assume a high test coverage, the less-frequent-testing strategies are broadly outperformed by their more-frequent counterparts under low test coverage (Supplementary Figures S3 and S4).

Based on the hospitalization risks at the two treatment initiation time ranges considered in the Phase 2/3 EPIC-HR trial, we fitted RRs and assumed a linear decline in efficacy to estimate the treatment efficacy levels associated with nirmatrelvir treatment across a range of days since symptom onset. To vary these assumptions, we considered trends that could capture two different time windows of efficacy beyond the range considered in the trial, and found little to no changes in our estimated RRs (Figure 2). We further note that while non-linear trends may marginally alter the magnitude of our expected RRs, with RRs inflated towards 1 if we assume a curvilinear decline consistent with a shorter efficacy window, they are unlikely to change the observed relative magnitude across strategies.

**Figure 2.**
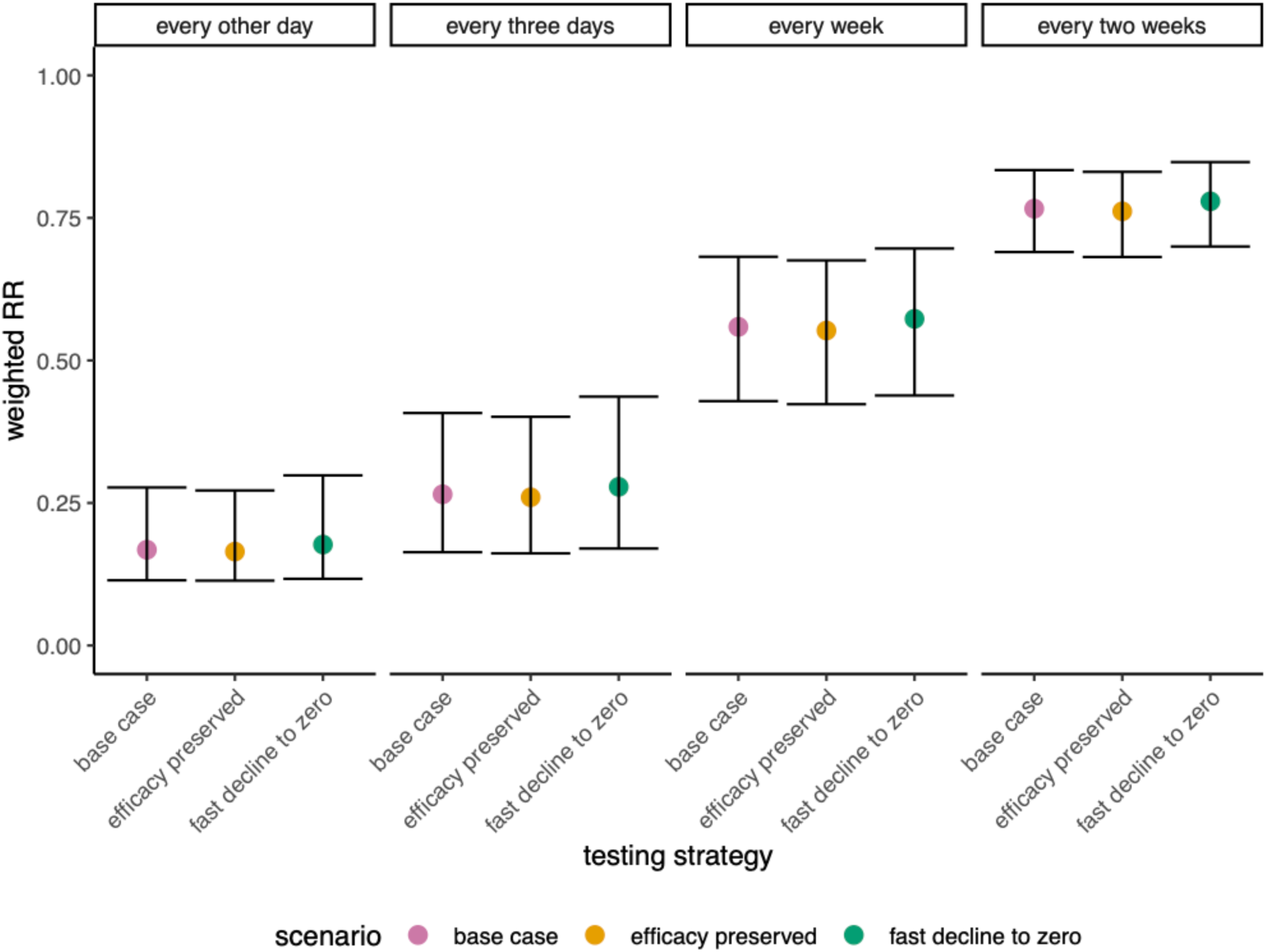
Median estimated weighted RRs of hospitalization by assumed treatment efficacy scenario (base case scenario, scenario with preserved efficacy following five days after symptom onset, and scenario with efficacy dropping to zero following days after symptom onset) across testing strategies: every other day, every three days, every week, and every two weeks. In all cases no positive-test-to-treatment delay and full test coverage were assumed. We do not assess the additional efficacy scenarios for the one-time testing strategy, because under this strategy, individuals who test positive take treatment on the day of testing; as such, the assumed RR trends beyond zero days since symptom onset are irrelevant.

**Figure 3.**
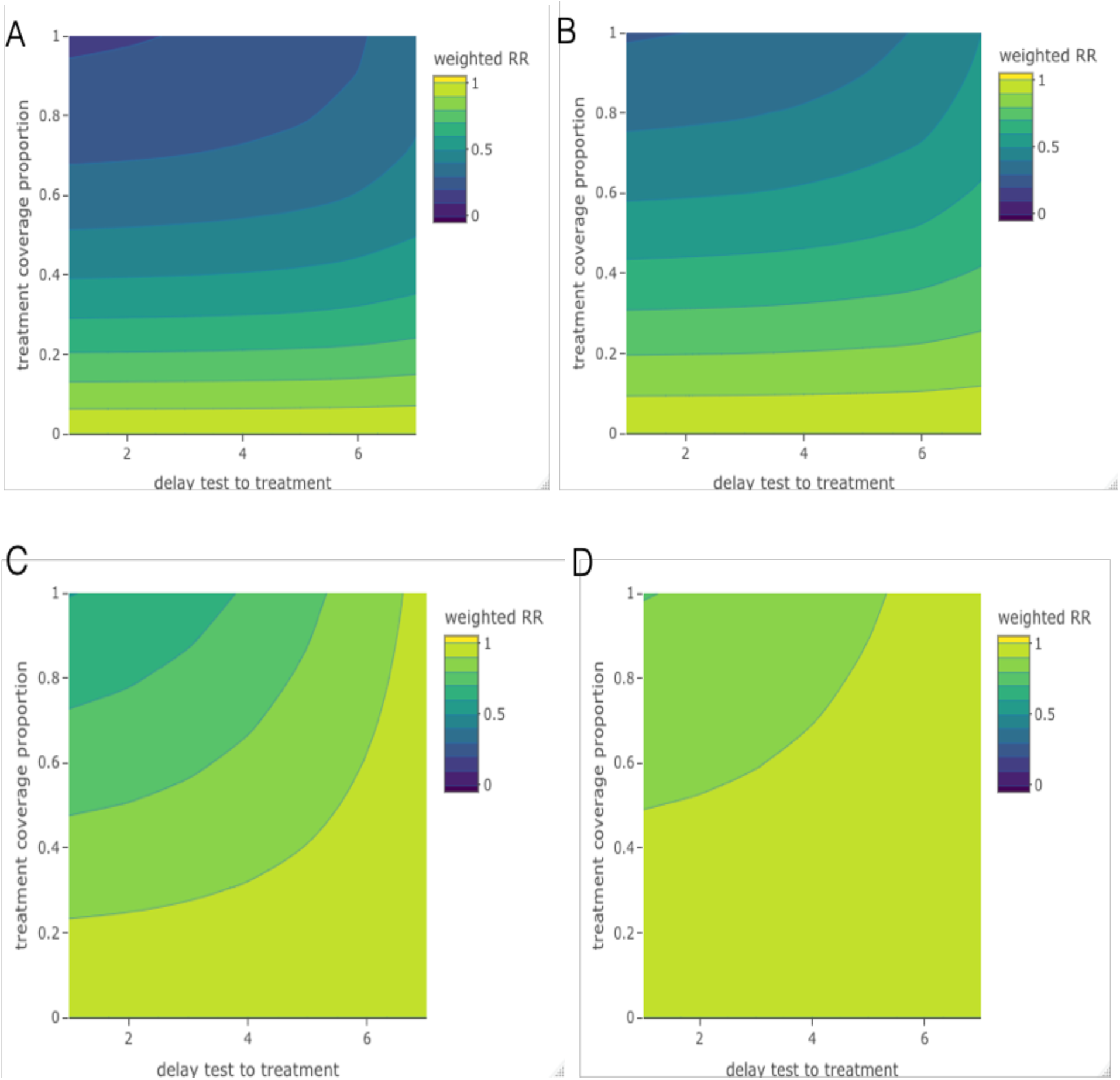
Sensitivity of estimated weighted RRs of hospitalization to positive-test-to-treatment delays (x-axis) up to seven days and treatment coverage proportions (y-axis) up to full coverage, by testing strategy (A-every other day, B-every three days, C-every week, D-every two weeks). Darker colors indicate lower weighted RRs, i.e. greater treatment-associated reductions in hospitalization in risks.

The positivity data from the Hellewell et al. analysis assumed an “LFT-like” cycle threshold (CT) of 28.^9^ If a lower CT threshold were used, we would expect the estimated RRs of hospitalization to increase and the corresponding proportion benefiting from the treatment to decrease, with the converse holding true for a higher assumed CT threshold. However, the ordering of RRs across strategies would once again persist. Patient data were collected in early 2020, such that time-specific positivity estimates were obtained from wild-type infections, with trends that could differ from the currently predominant variant.^9^ In contrast, hospitalization risks were estimated using data from July 2021 and thus likely were recorded on largely Delta-infected patients.^5^ We note that if the prevailing variant were associated with a substantially increased or reduced risk of hospitalization, this would likely hold true for patients in general, regardless of whether they received treatment, such that the relative risks of hospitalization would remain relatively unchanged. If, however, treatment is effective for a longer (or shorter) period of time, we would observe a narrowing (or expanding) benefit of more periodic testing. Additionally, if treatment were found to be effective under the same time frame, but to a greater extent, we would anticipate increased benefits under all testing strategies. Thus, it is important to update our results specific to the current variant and among vaccinated populations^5^, once new data become available.

Despite the promising role of rapid testing that we observe here, it is important to acknowledge the costs that result from such testing policies. For instance, approximately six billion pounds have been paid by the UK government for their mass lateral flow distribution plan, which concluded on April 1, 2022.^15^ However, we note that under a focused testing plan, prioritizing frequent testing among those who are most likely to be prescribed treatment upon a positive test, as is the subject of attention here, these costs would be considerably less. From the perspective of patient populations^16^, costs include those associated with (highly unlikely^17^) false positive results, such as missed earnings from work, missed medical appointments for other health conditions, and stress-related mental health consequences.^18^ However, with a substantial proportion of the population successfully being linked to treatment due to testing, there may be significant cost savings (to both hospitals and patients) from averted hospitalizations, specifically among patients who may be driven to debt as a result of these expenses.^19–21^

In sum, we characterized how rapid testing may facilitate treatment benefits among those most likely to be hospitalized, with more frequent testing yielding the best results. While we also observed notable benefits under a one-time test policy, this regime requires that individuals recognise symptoms and, as with the other strategies, have tests available to use soon after symptoms emerge. Test and treatment access matters: high coverage and short delays from testing to treatment are necessary to achieve large benefits. Spatially-refined testing strategies might further support disadvantaged communities where vulnerabilities to severe disease and barriers to testing and treatment are most concentrated. Finally, regular testing is potentially cost-saving, particularly in high-prevalence settings, as it is associated with dramatically reducing hospitalizations, which may outweigh the costs of testing and treatment distribution.

## Data Availability

All code is available at https://github.com/goshgondar2018/LFT_treatment_analysis

## Acknowledgments

TFM acknowledges funding from the Harvard Center for Communicable Disease Dynamics’ Program Support fund and the Harvard T.H. Chan School of Public Health’s Rose Traveling Fellowship. CAD acknowledges support from the MRC Centre for Global Infectious Disease Analysis, the NIHR Health Protection Research Unit in Emerging and Zoonotic Infections and the NIHR-funded Vaccine Efficacy Evaluation for Priority Emerging Diseases (PR-OD-1017-20007).

The computations in this paper were run on the FASRC Cannon cluster supported by the FAS Division of Science Research Computing Group at Harvard University.

