## Supplementary Appendix for "The impact of repeated rapid test strategies on the effectiveness of at-home antiviral treatments for SARS-CoV-2"

Our analysis required two primary sources of data to estimate probability-weighted risk ratios (RRs): positivity curves and treatment efficacy-associated RRs. To parameterize the test positive probabilities, we used an estimated lateral flow test "(LFT)-like" positivity curve generated from a Hellewell et al. Bayesian model, which reports the posterior probability of testing positive for each day since infection from 0 to 30 days, across 4000 Markov chain Monte Carlo (MCMC) draws.[1] That is, while the patient population in this analysis were routinely administered PCR - and not antigen - tests, the authors provided two scenarios estimating expected test positive probabilities under antigen testing, by defining lower CT thresholds that would better reflect the diagnostic ability of LFTs. To parameterize RRs of hospitalization, we used data reported in the latest EPIC-HR press release, namely the number of hospitalized subjects in the treatment and placebo groups, and the total size of each treatment arm, stratified by when treatment was initiated, i.e. within three or five days since symptom onset, in a trial population of SARS-CoV-2-positive patients at an elevated risk of severe disease.[2,3]

### A. Estimating weighted RRs, proportion given and benefiting from the treatment

To estimate RRs over a range of days since symptom onset, we fit a log-binomial model to the reported hospitalization counts by assigning a series of treatment-labeled 1s and 0s to reflect the recorded hospitalizations and non-hospitalizations within each arm, with covariates representing days since symptom onset and treatment group. We assumed a linear relationship between the continuous predictors (day and the treatment x day interaction) and the log risk of hospitalization as well as an additive effect of treatment. From our model fit, we extracted RR estimates for each day on the range [0,7] days since symptom onset and defined a RR of 1 (i.e. no treatment effect) thereafter, given the limited, if any, efficacy that is expected to occur a week following symptom onset. We additionally fit a logistic model to determine whether our results were sensitive to whether RRs and ORs were employed, and observed nearly equivalent results, as expected given the rarity of the hospitalization outcome (Figure S1).

Consistent with the method detailed in Hellewell et al. for estimating probabilities of pre-symptom onset symptomatic detection[1], assuming a CT threshold of 28, we used the probability of either first testing positive at the testing day $t_{max}$ (the product of the test negative probabilities for each test day up until day $t_{max}$ and the test positive probability at day $t_{max}$) to weight, among the treated, the corresponding RR at day $t_{max}$, *or* testing negative at all testing days, weighted by an RR of 1 corresponding to no treatment; we then averaged these weighted RRs over all possible testing sequences, as shown in Equation 1. We replicated this process over the 4000 simulations of positivity data and for each testing strategy (every other day, every three days, once a week, and once every two weeks). To summarize our results for each strategy, we extracted median weighted RRs and an accompanying 95% confidence interval (CI). To match hospitalization risks reported with the time reference of days since symptom onset to test positive probabilities reported with the time reference of days since infection, we sampled incubation periods (the times from infection to symptom onset) - assuming a lognormal distribution consistent with pooled estimates from a McAloon et al. meta-analysis[4]- at each iteration and used the rounded incubation periods to index time relative to symptom onset rather than infection and identify the appropriate RR for each day. We allowed for possible delays in x days from testing positive to treatment by additionally setting a variable which shifts this period by x days. In our main analyses, we assumed a zero-day positive-test-to-treatment delay corresponding to potential plans to distribute antiviral pills to households to store for immediate use if necessary and further assumed full treatment coverage.

$$Weighted\ RR_{strategy\ ts} = \frac{1}{index}\{\textstyle\sum_{seq\in tseq}(\prod_{t}^{t_{max}-1} Pr(test-)_t * Pr(test+)_{t_{max}} * RR_{t_{max}+delay-incubation.period} * P_{tmnt}) + \displaystyle\sum_{i=0}^{index}(\prod_{i,i+index,\ldots,30-index} Pr(test-)_{t2} * 1)\}$$

[Equation 1]

where

$seq \in tseq$ denotes a given testing sequence seq - each defined by when testing is initiated - in the full set of possible sequences consistent with strategy ts,

t is initialized as the first time point for each testing sequence,

$t_{max}$ is the final day of that testing sequence,

$RR_{t_{max}+delay-incubation.period}$ denotes the risk ratio associated with the final day of that testing sequence, back-shifted for the sampled incubation period, adding any test-to-treatment delays

$P_{tmnt}$ denotes the the treatment proportion,

index = 2, 3, 7,and 14 for every other day, every three days, once a week, and once every two weeks, respectively

We subsequently estimated the proportion of the infected population who could be offered the treatment (which can be interpreted as "given the treatment" under full treatment coverage and uptake), representing all infected individuals who test positive at any point. Specifically, this proportion can be expressed as the complement of the mean across testing sequences of the probability of testing negative at all time points, which captures all always-test-negative possibilities, standardized by the number of possible sequences, as described in Equation 2 below.

$$Proportion\ offered\ treatment_{strategy\ ts} = 1 - (mean_{[0,index]} \textstyle\prod_{i,i+index,\ldots,30-index} Pr(test-)_{t2})$$

[Equation 2]

Finally, we estimated the proportion of the infected population who derive some benefit from treatment, that is, those able to initiate treatment during a period when the RR does not equal 1. To do so, for each possible testing sequence, we first assigned an indicator variable equal to 1 for the positive testing time if it precedes the day when RR = 1 and equal to 0 otherwise (which incorporates those never testing positive and those testing positive too late to benefit from treatment). We then averaged test-positive-probability-weighted indicators across all testing sequences to give the mean proportion benefiting from the treatment. $I(day_i <= 7 + incubation.period)$ denotes whether the testing day plus any test-to-treatment delay falls within seven days of the incubation period (that is, seven days following symptom onset), when we assume treatment is no longer efficacious. To account for imperfect testing, we multiplicatively reduced our estimated proportions benefiting from treatment by the proportion testing.

$$Proportion\ benefiting_{strategy\ ts} =$$

$$P_{test} * \frac{1}{index}\{\sum_{seq \in tseq} (\prod_{t}^{t_{max}-1} Pr(test-)_t * \mathrm{Pr}(test+)_{t_{max}} * (I(day_i <= 7 + incubation.period)))$$

[Equation 3]

where $P_{test}$ denotes the testing proportion.

In the context of a strategy of testing once only after once symptoms develop, assuming zero-day positive-test-to-treatment delays, individuals will always take treatment on the day of their (positive) test result and thus, 1) all those who are offered treatment benefit from it and 2) there is only one possible testing probability, with a corresponding risk ratio of hospitalization that is always that of zero days since symptom onset. As before, we report medians and a corresponding 95% CI to summarize estimates across the 4000 samples.

### B. Sensitivity Analyses

To accommodate shorter incubation periods that have been observed among Omicron infections, we considered a Weibull incubation period distribution found to generate the optimal fit to symptom onset times in an analysis of Omicron-infected subjects in Norway.[5] Our outcomes of focus in this sensitivity analysis are weighted RRs and proportions benefited under each testing strategy (Supplementary Figure S2), as the estimated proportions offered treatment, which not depend on when an individual tests positive, will remain unchanged.

We additionally evaluated how varying our assumed period of treatment efficacy, reflecting waning of treatment impacts, would influence the marginal benefits of more frequent rapid testing. In addition to our base case scenario, where we assumed a linear decline in efficacy, as described previously, we considered two additional scenarios assuming either 1) the RR consistent with initiation at five days since symptom onset is preserved following five days after symptom onset until the latest positive test (referred to as 'efficacy preserved' in Figure 2) or 2) the RR goes to one, reflecting zero efficacy, after five days since symptom onset (referred to as 'fast decline to zero' in Figure 2). Weighted RRs are estimated for each scenario (Figure 2).

We subsequently considered the combined impacts of treatment coverage and positive-test-to-treatment delays on weighted RRs by jointly varying the treatment coverage proportion across the range [0,1] by 0.1 (10%) unit increments and the delay from testing positive to treatment across the range [0,7] days, under our main treatment efficacy scenario. For this, we used median estimated day-specific test positivity values. We then compared weighted RRs across all resulting combinations (Figure 3). As described previously, in a separate analysis, we examined the isolated impacts of test access on proportions offered and benefiting from treatment through a series of scenarios in which we varied test coverage across a range of values from 0 to 1 (Supplementary Figures S3 and S4).

All code is available at: https://github.com/goshgondar2018/LFT_treatment_analysis

Figures

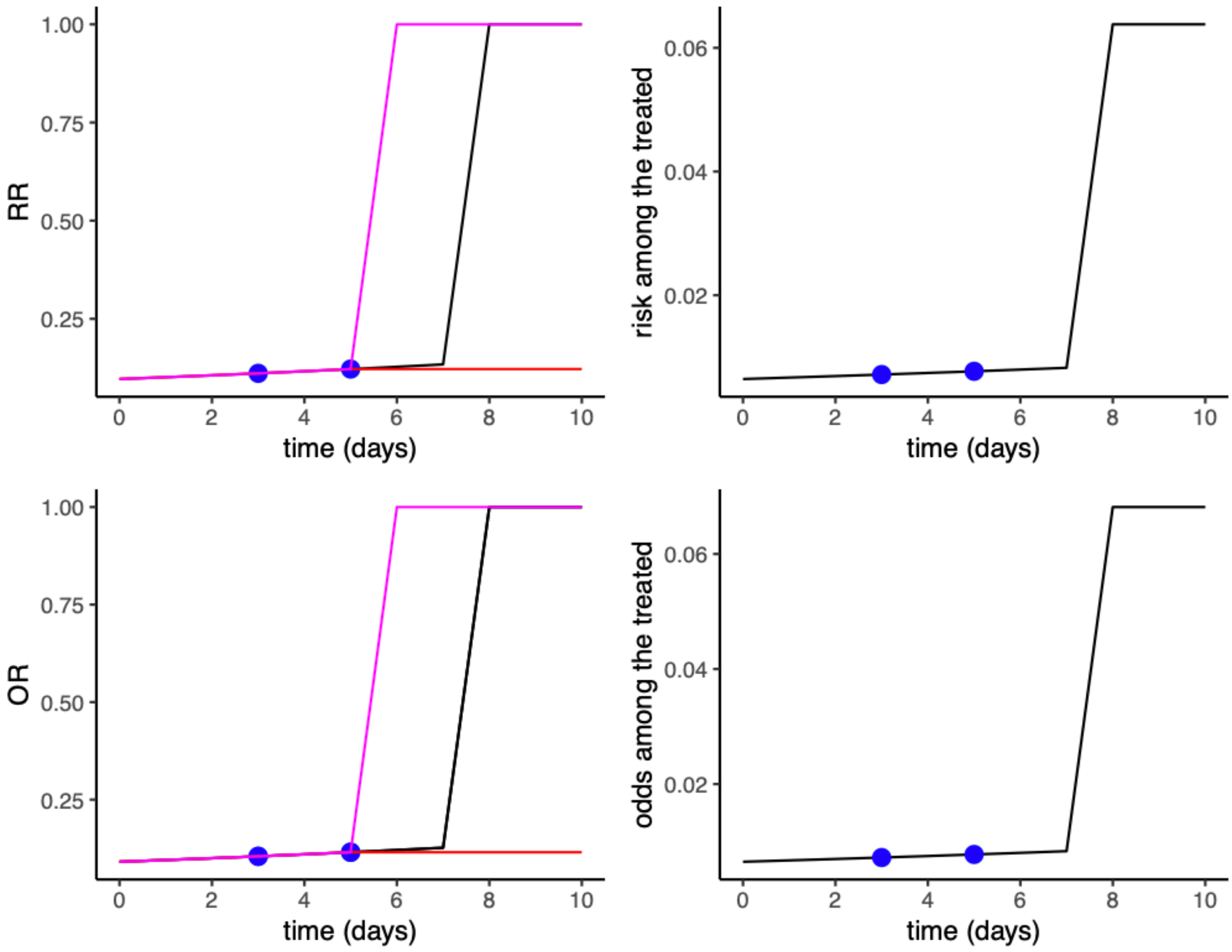


**Supplementary Figure S1.** (A) Estimated relative risks of hospitalization as a function of time of treatment relative to symptom onset (B) Estimated risks of hospitalization among subjects treated with nirmatrelvir as a function of time of treatment relative to symptom onset (C) Estimated odds ratio of hospitalization as a function of time of treatment relative to symptom onset (D) Estimated odds of hospitalization among subjects treated with nirmatrelvir as a function of time of treatment relative to symptom onset. Blue points indicate observed RRs (A), risks among the treated (B), ORs (C) and odds among the treated (D) at three and five days of symptom onset as reported in the EPIC-HR press release[2]. In panels A and C, the black lines capture estimates under our main scenario, the pink lines capture estimates under the fast decline to zero efficacy scenario, and the red lines capture estimates under our efficacy preserved scenario.

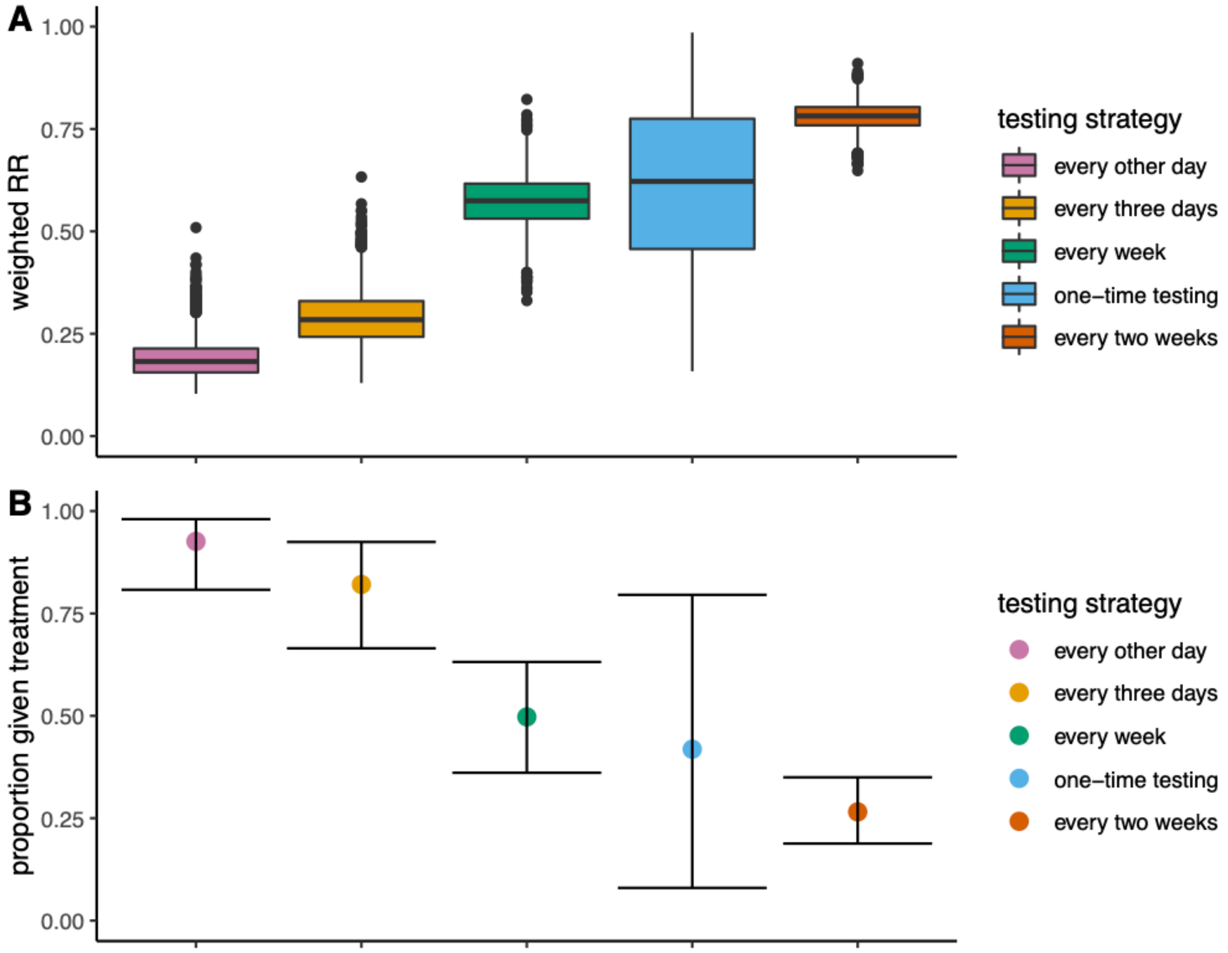


**Supplementary Figure S2.** Replicate of Figure 1, with results from the shorter incubation period scenario (A) Distribution of estimated weighted risk ratios of hospitalization by testing strategy: every other day, every three days, every week, one-time testing and every two weeks. Medians are marked by solid horizontal lines, each box includes the full interquartile range, and plotted points are those which extend beyond the upper/lower quartile +/- 1.5*interquartile range (B) Estimated proportions given treatment by testing strategy (including the one-time post-symptom-onset testing strategy) with 95% uncertainty intervals. In all cases no positive-test-to-treatment delay and full treatment and test coverage were assumed.

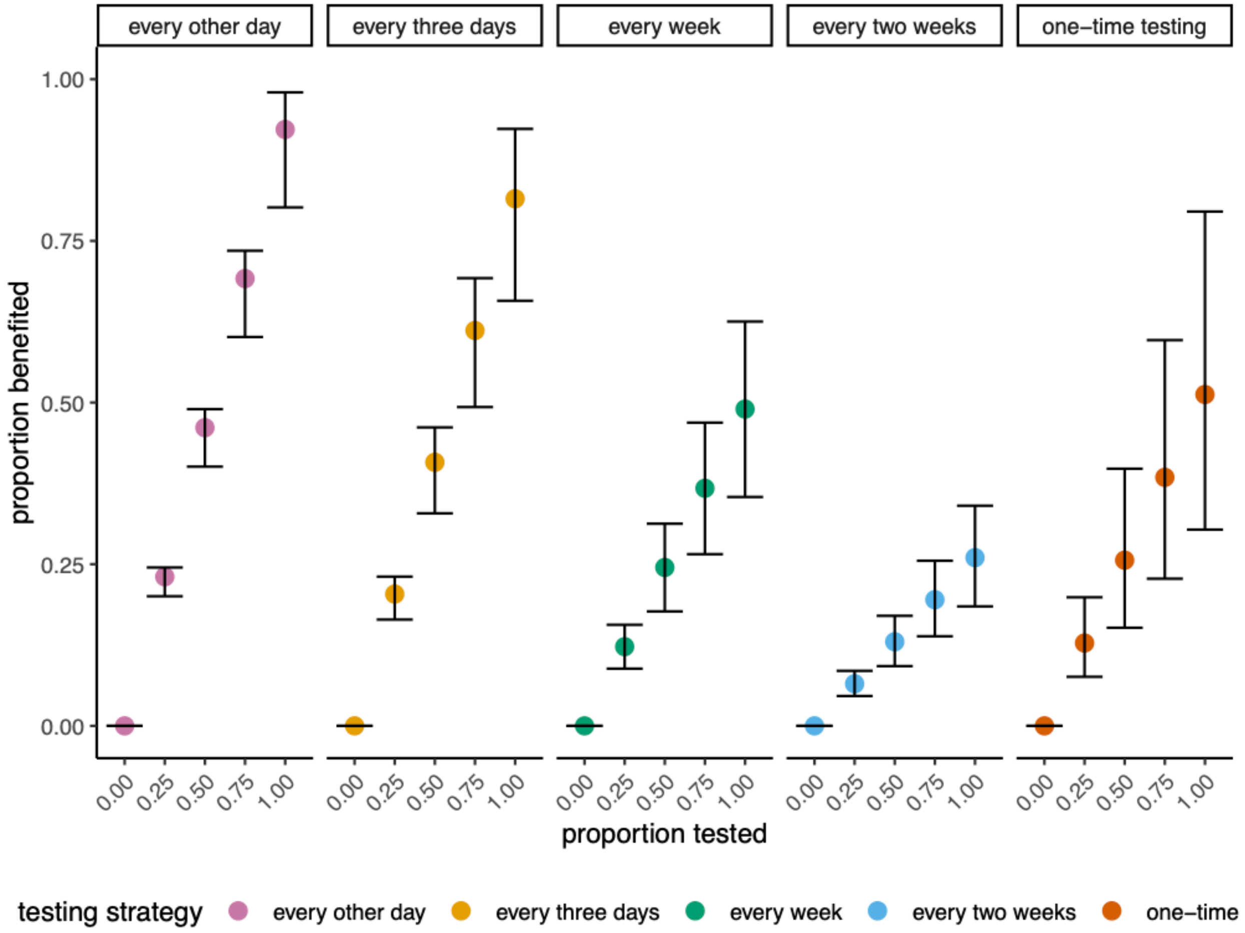


**Supplementary Figure S3.** Estimated proportions who benefit from treatment, with corresponding 95% CIs, as a function of proportion of the population who tests. Each vertical panel corresponds to a testing strategy.

| **Testing Strategy** | **Median RR (95% CI) [baseline incubation period scenario]** | **Median RR (95% CI) [shorter incubation period scenario]** | **Median proportion benefited (95 % CI) [baseline incubation period scenario]** | **Median proportion benefited (95 % CI) [shorter incubation period scenario]** |
|---|---|---|---|---|
| **Every other day** | 0.168 (0.114-0.277) | 0.182 (0.122-0.299) | 0.922 (0.802-0.980) | 0.920 (0.799-0.979) |
| **Every three days** | 0.265 (0.164-0.408) | 0.284 (0.175-0.431) | 0.815 (0.657-0.923) | 0.812 (0.654-0.922) |
| **Once every week** | 0.559 (0.429-0.682) | 0.575 (0.447-0.695) | 0.490 (0.354-0.625) | 0.487 (0.351-0.624) |
| **Once every two weeks** | 0.766 (0.690-0.834) | 0.782 (0.712-0.844) | 0.260 (0.185-0.340) | 0.257 (0.183-0.336) |
| **One-time testing** | 0.537 (0.282-0.726) | 0.622 (0.281-0.928) | 0.512 (0.303-0.795) | 0.418 (0.0798-0.795) |

**Supplementary Table S1**. Estimated weighted risk ratios of hospitalization, under the incubation period distribution assumed in our main analysis (baseline) and in our shorter incubation period scenario
